# Inequalities in Mental Health: Age-related Trends Across Pandemic Phases in Singapore

**DOI:** 10.1101/2024.08.25.24312468

**Authors:** Nicole Shu En Goh, Christian Morgenstern, Alex Jie Wei Yap, Stanford Chun Yin Wong, Mythily Subramaniam, Edimansyah Bin Abdin, David Chien Boon Lye, Kelvin Bryan Tan, Sharon Hui Xuan Tan

## Abstract

**Background:** In the sphere of mental health, little population wide-scale research has been done in Asia on the pandemic’s differential impacts on different sociodemographic groups over time.

**Methods:** This study evaluates symptoms of anxiety and depression within different age groups in Singapore across different pandemic phases. Symptoms of depression and anxiety were measured using the Patient Health Questionnaire-4 (PHQ-4), in which higher scores indicate more symptoms and lower scores indicate less. Cross-sectional data from 34,429 individuals aged 18 years and above in Singapore between 27 April 2020 and 27 September 2021 were analyzed. Multivariable logistic and linear regression were performed on PHQ-4 scores over pandemic phases and by age.

**Findings:** Overall symptoms of depression and anxiety did not vary significantly across pandemic phases in Singapore. However, compared to Circuit Breaker, younger age groups reported higher PHQ-4 scores as restrictions lifted (ages 18-29: *beta (*β*)=0·59, 95% CI [0·22, 0·97]*; ages 30-39: β*=0·44, 95% CI [0·08, 0·80])*, while older adults reported lower scores (ages 50-59: β*=-0·45, 95% CI [-0·80, -0·10];* ages 60 and above: β*=-0·38, 95% CI [-0·73, -0·02])*. This was associated with more symptoms of anxiety among younger adults, and less symptoms of depression and anxiety among older adults.

**Interpretation:** In Singapore, psychological responses to pandemic restrictions may be heterogenous across different age groups and across time. The study can inform policymakers’ decisions in distributing mental health resources during a crisis.

**Funding:** The funders had no role in study design, data collection, data analysis, data interpretation, or writing of the report.

## 1. Introduction

While most countries have transitioned to a “return to normal” phase in the management of COVID-19^1^, the long-term physical, psychological, and economic impacts of the COVID-19 pandemic remain the subject of much research and concern. Country-wide lockdowns during COVID-19 created social distance within communities and nations in a previously interconnected global environment, contributing to spikes in levels of depression and anxiety across the world^2–4^.

The COVID-19 pandemic had widespread impacts on mental well-being on populations worldwide. Globally, there was a high prevalence of psychological distress during COVID-19^5–8^. While lockdown measures were effective in reducing transmission and mortality^9^, they were associated with risk factors for poorer psychological well-being, such as increased feelings of isolation and more frequently reported feelings of hopelessness and worry^9-13^. Consistent with these findings, research in Denmark and the United States (US) found that population symptoms of anxiety and depression were worst during lockdown phases and improved when restrictions were relaxed^4, 14^.

Many high-income Asian countries such as Singapore, Japan, and South Korea employed a different policy strategy than Western countries such as US and Denmark. Where US and Denmark employed a mitigation strategy, aiming only to limit the effects of COVID-19 within their countries, these Asian countries employed an elimination strategy, which aimed to eliminate community transmission of COVID-19 within their borders^9, 15^. National restriction timings and intensities tended to differ with their policy strategy: those using an elimination strategy generally had more rigorous policies at the beginning of the pandemic, but relaxed these restrictions and became less stringent later than countries pursuing a mitigation strategy^9^. However, no research exists in Asia on how symptoms of anxiety and depression changed across COVID restriction periods, particularly within different sociodemographic groups. It is possible that, in a high-income Asian country like Singapore, relaxation of restrictions led to greater reduction in psychological symptoms reported than in US or Denmark, given the better health and mortality outcomes associated with an elimination strategy than a mitigation one^9, 15^. However, it is also possible that relaxation of restrictions might create more anxiety within high-income Asian countries as community cases increase, as this runs counter to the national goal of eliminating community transmission.

Either way, these differences in population response to relaxing restrictions could also exacerbate differences in psychological symptoms between sociodemographic groups. As such, while research in US and Denmark found that trends in depression and anxiety symptoms reported over time were largely similar across sociodemographic groups^4, 14^, differentiation in subgroup trends might be observed in a country such as Singapore.

In particular, age may affect individual psychological recovery from the pandemic over time. During COVID-19, younger adults reported worse psychological impact than older adults from stressors such as lockdown^16^, but more improvement in mood from positive experiences as well^17^. On the other hand, older age was associated with increased emotional resilience and well-being at the baseline^18,19^, but also with slower recovery that put them at higher risk of post-COVID depression or anxiety upon requiring acute care for COVID-19^20^. Taken together, literature suggests that younger adults may suffer more symptoms of anxiety and depression during lockdowns and the earlier stages of the pandemic but experience faster recovery from symptoms as restrictions are lifted. However, it is unknown as whether these trends persist throughout the pandemic, particularly in an Asian context such as in Singapore.

Using repeated cross-sectional data, this study investigated the differential impacts of the pandemic on depression and anxiety symptoms of Singapore’s population and its different age groups, both during the initial lockdown period and as the pandemic progressed.

### Research in context

#### Evidence before this study

We searched Google Scholar for articles published in English using the search terms “Asia”, “COVID”, “age” or “age group”, and “mental health trends”. Our search identified over a million articles published between 1^st^ January 2020 to the present—none of which, to our knowledge, analysed psychological symptoms among age groups across restriction periods in a high-income Asian country. Previous studies were conducted primarily in European or American contexts or were limited to smaller population subgroups or restricted time periods, rendering it difficult to track trends by age groups within these countries. Few studies compared trends in mental health symptoms within age groups across time and with restrictions.

#### Added value of this study

This study shows that trends in symptoms of depression and anxiety in high-income Asian countries like Singapore may be heterogenous across age groups. Younger adults report more symptoms at the baseline and tend to report more symptoms across the pandemic despite relaxation of restrictions, while older adults report fewer symptoms at the baseline and tend to report symptoms less frequently as restrictions are lifted. As such, inequalities in mental health between age groups appear to worsen across the pandemic.

#### Implications of all the available evidence

The study’s findings support existing literature documenting fewer affective symptoms and greater resilience among older adults in the face of negative experiences. Despite older adults experiencing greater health risks during the pandemic, psychological symptoms among younger adults appear to increase disproportionately as the pandemic goes on. Increased psychological support should be allocated to younger adults during epidemics and crises. Psychological impacts of the pandemic may present differently in older adults, such as through physiological rather than affective symptoms, and further research is required.

## 2. Methods and Measures

### 2.1. Participants

Data from Singapore were drawn from the COVID-19 Behavioral Tracker^21^, a repeated cross-sectional online survey launched across 30 countries in a collaboration between YouGov and Imperial College London’s Institute of Global Health Innovation.

In Singapore, the behavioral tracker was sent out in 38 waves from 3 April 2020 to 27 September 2021, with a total of 38,000 adult respondents. The PHQ-4 questionnaire was administered to participants in the latter 36 waves. Data from these 36 waves was analyzed, comprising 34,429 responses collected between 27 April 2020 and 27 September 2021 (Supplementary Appendix Table S1).

### 2.2. Pandemic Restrictions

Participants’ responses were grouped into one of six periods according to the restrictions in place on the date they submitted their survey (Figure 1). The restriction periods were as follows: Firstly, the Circuit Breaker, when all non-essential services were suspended. Secondly, Phase 1 and Phase 2, which allowed for some visiting of other family households, gatherings of up to five people, and dining-in at food and beverage (F&B) outlets. Third, Phase 3, which allowed for social gatherings of up to eight people and for attractions to operate at higher capacities (65% capacity, rather than 50%). Fourth, Heightened Alert, where group sizes were scaled back to five people and vaccine-differentiated dine-in measures were introduced (vaccinated and unvaccinated individuals were allowed to dine in in groups of five and two respectively). Fifth, Transition Stage, where fully vaccinated individuals were allowed to participate in higher-risk activities with their masks removed and people were allowed to dine-in at hawker centers in groups of two. And sixth, when travel restrictions were relaxed slightly and travelers from certain countries were no longer required to quarantine when visiting Singapore or to quarantine in their own homes (Figure 1 and Supplementary Appendix Table S8).

The Circuit Breaker was selected as the study baseline. Population characteristics were similar across all restriction periods (Supplementary Appendix Table S6), although the mean PHQ-4 scores of 18-to 29-year-olds increased from 4·40 (SD=3·02) to 5·01 (SD=3·12) from Circuit Breaker to when border restrictions were relaxed, and those of 30-to 39-year-olds increased from 3·98 (SD=3·10) to 4·36 (SD=3·21) in the same time frame (Supplementary Appendix Table S7).

### 2.3. Psychological Distress

Psychological distress was measured by participants’ total scores on the Patient Health Questionnaire-4 (PHQ-4), a screening tool for anxiety and depression that has been validated in Singapore^22, 23^. The PHQ-4 is a four-item questionnaire made up of the PHQ-2, which measures diminished pleasure and feelings of depression over the last two weeks, and the General Anxiety Questionnaire-2 (GAD-2), which measures feelings of anxiety and uncontrollable worrying over the last two weeks. Each question is measured on a four-point Likert scale: 0 (“Not at all”), 1 (“Several days”), 2 (“More than half the days”), and 3 (“Nearly every day”). Total scores on the PHQ-4 therefore range from 0 to 12, with higher scores indicating that an individual demonstrates more symptoms of anxiety and depression. Whether or not a person exhibits significant depressive symptoms and symptoms of anxiety is measured by whether they score above three in the PHQ-2 and GAD-2 respectively^24^.

### 2.4. Covariates and statistical analysis

Self-reported data on sex (male, female), mental health condition diagnosis (yes, no, prefer not to say), age, employment status, and vaccination availability were also captured. Age was grouped evenly into five bands (18-29, 30-39, 40-49, 50-59, and 60+ years of age), while vaccination availability was classified into two (vaccine available and other). For vaccination availability, “other” was used as a category to encompass both responses of “the vaccine is not available to me” and non-responses, as the question on vaccine availability was only introduced after vaccines were rolled out. As such, a non-response could indicate either that no vaccine was available to the population or that a participant preferred not to disclose their response. Employment status was classified into four groups (full time employment, unemployed, not working, other), with students and part-timers classified under “other” and retirees classified under “not working”.

The analyses used sample weights based on age, gender, and region within country to produce nationally representative estimates. Univariate and multivariable linear and logistic regressions were used to analyze the effect of period on symptoms of anxiety and depression (as measured by participants’ total PHQ-4 scores) across the entire study population. Subgroup analyses were carried out for each age group. Subsequently, multivariable logistic regressions were used to analyze the prevalence of depressive symptoms and symptoms of anxiety within age groups over time. All analyses adjusted for the confounding effects of age band, gender, employment status, existing mental health condition, and reported availability of vaccination.

## 3. Results

The demographic characteristics of the survey population, pooled across all 36 waves of the YouGov survey in which the PHQ4 was administered, are presented in Table 1. Younger adults experienced more symptoms of depression and anxiety than older adults, with the youngest age band (ages 18 to 29) scoring the highest on the PHQ-4 (*Mean=4·71, SD=3·17*) and the oldest age band of those over 60 scoring the lowest (*Mean=2·06, SD=2·90*). Those with mental health conditions *(Mean=6·06, SD=3·77)*, as well as those who were unemployed (*Mean=4·57, SD=3·72*), reported levels of psychological symptoms that were higher than the population average (*Mean=3·51, SD=3·27*).

### 3.1. Trends in psychological distress

Across participants, there was no statistically significant difference in overall symptoms of depression and anxiety throughout the pandemic (Table 2). Within age groups, however, trends in PHQ-4 scores differed across time (Table 2). As restrictions were lifted, younger adults tended to report more anxiety and depression symptoms, while older adults tended to report fewer. Compared with during the Circuit Breaker, those aged 18 to 29 reported depression and anxiety symptoms more often during Heightened Alert (β*=0·40, 95% CI [0·08,0·73])*, and Relaxed Border Restrictions (β*=0·59, 95% CI [0·22, 0·97])* periods, while those aged 30 to 39 reported symptoms more often during Phase 3 (β*=0·40, 95% CI [0·08, 0·71]*) and Relaxed Border Restrictions (β*=0·44, 95% CI [0·08, 0·80])* stages. Meanwhile, compared with during Circuit Breaker, those aged 50 to 59 reported anxiety and depression symptoms less often during Phase 1 and 2 (β*=-0·33, 95% CI [-0·60, -0·06])*, Phase 3 (β*=-0·54, 95% CI = [-0·85, -0·22])*, Heightened Alert (β*=-0·32, 95% CI [-0·61, -0·02])*, and Relaxed Border Restrictions (β*=-0·45, 95% CI [-0·80, -0·10])* stages, while those over 60 reported symptoms less often during Phase 3 *(−0·39, 95% CI [-0·72, 0·07])*, Heightened Alert *(−0·44, 95% CI = [-0·75, -0·14)]*, Transition (β*=-0·53, 95% CI [-1·03, -0·04])*, and Relaxed Border Restrictions (β*=-0·38, 95% CI [-0·73, -0·02])* stages.

Trends in symptoms of depression and anxiety were analyzed for other population subgroups as well (Supplementary Appendix Table S8). In general, the number of symptoms reported by these subgroups did not change significantly following the Circuit Breaker. There was a slight fall in symptoms reported by females (*OR=-0·21, 95% CI [-0·40, -0·02]*) and individuals who were not working *(OR=-0·40, 95% CI [-0·79, -0·02])* from Circuit Breaker to the Heightened Alert stage, but these trends did not persist when Singapore began to reopen its economy more fully (during its Transition stage and when it relaxed its border restrictions).

### 3.2. Trends in significant symptoms of depression and anxiety

Population anxiety symptoms, as measured by scoring three or higher on the GAD-2, did not differ in a statistically significant way across the pandemic. No statistically significant difference in the prevalence of symptoms reported was found when comparing between the Circuit Breaker and any other restriction period (Table 3). However, sub-group analyses found that younger adults were more likely to report symptoms of anxiety when restrictions were lifted. Those aged 18 to 29 and 30 to 39 were 1·48 *(95% CI [1·15, 1·91])* and 1·29 *(95% CI [1·00, 1·66])* times more likely to report symptoms of anxiety respectively when border restrictions were relaxed compared with during Circuit Breaker. Older adults, however, were less likely to report anxiety symptoms over time, with those aged 60 and above being 0·60 *(95% CI [0·43, 0·83])* times less likely to exhibit symptoms of anxiety when border restrictions were relaxed compared with during Circuit Breaker.

For the whole population, fewer depressive symptoms were reported in every other restriction period compared with Circuit Breaker (Table 4), which may be related to the lower likelihood of older adults to report symptoms later in the pandemic. In particular, those aged 50 to 59 were less likely to report depressive symptoms during Phase 1 (*OR=0·75, 95% CI [0·61, 0·93])*, Phase 3 *(OR=0·63, 95% CI [0·49, 0·81])*, Heightened Alert stages *(OR=0·78, 95% CI [0·62, 0·98])* and when border restrictions were relaxed *(OR=0·64, 95% CI [0·48, 0·85])* than during Circuit Breaker. Similarly, compared with during Circuit Breaker, those aged above 60 were less likely to report depressive symptoms during Phase 3 *(OR=0·73, 95% CI [0·54, 0·98])*, Heightened Alert *(OR=0·69, 95% CI [0·52, 0·92])* and Transition stages *(OR=0·48, 95% CI [0·28, 0·83])*, and when border restrictions were relaxed (*OR=0·64, 95% CI [0·45, 0·91]*). Among younger adults (ages 49 and younger), no statistically significant change in depressive symptoms reported was observed throughout the pandemic.

## 4. Discussion

This paper is one of few detailing how trends in anxiety and depression symptoms differ by sociodemographic groups in the COVID-19 pandemic. It is one of the first studies to analyze COVID-19 mental health trends across multiple time points, particularly in a developed Asian country such as Singapore. This is crucial for identifying vulnerable populations in future crises or pandemics, as well as for evaluating the adequacy of existing systems of psychological support for pandemic or crisis recovery. Findings suggest that the prevalence of such symptoms may not naturally return to baseline post-COVID, and that in certain population subgroups, symptoms of anxiety and depression may worsen in the face of relaxing restrictions and prolonged exposure to the pandemic.

For Singaporean adults, changes in anxiety symptoms appeared to contribute to changes in PHQ-4 scores, with older adults tending to report fewer symptoms and younger adults tending to report more symptoms over time. This supports research that has found age to be a moderator of anxiety in the face of COVID exposure^25^. Increased symptoms of anxiety among younger adults may be attributable to fears of job loss or among students, concerns regarding their ability to find a job. Pandemic restrictions and lockdowns led to increased business shutdowns and decreased economic activity^26^, which in turn were associated with income losses and fears of unemployment^27^. Despite increased economic activity as restrictions were lifted, young adults’ concerns regarding job security and the state of the economy may have persisted^28^. Older adults may have experienced less anxiety regarding these closures, as they may have already retired or were planning to do so soon. Furthermore, among students, these employment concerns may be compounded by academic anxiety or poorer academic performance due to challenges associated with distance learning, such as difficulties obtaining sufficient academic support and attenuated teacher-student relationships^29, 30^.

Contrary to expectations, social isolation in Singapore due to the pandemic may also have impacted younger adults more negatively than older ones. In the initial stages of the pandemic, when lockdown measures were first implemented, there were concerns that social isolation would create a mental health crisis among older adults. Indeed, isolation among the elderly was associated with poor behavioral and psychological outcomes^31^. However, isolation and loneliness are not limited to older adults, and literature suggests that younger adults may report feeling lonely more often than older ones^32–34^. Social isolation may be associated with worse health and behavioral outcomes among younger than older adults^35^. It is therefore possible that increased symptoms of anxiety and depression among younger adults compared with older ones may be the differential results of loneliness and isolation stemming from COVID-19 concerns and social distancing^36^.

Overall, the findings of this study suggest that, in Singapore, older adults may be less prone to exhibiting affective symptoms of depression and anxiety in the face of stressors and may experience a quicker reduction in these symptoms over time. In the case of COVID-19, it is possible that the pandemic caused fewer disruptions to the lives of older Singaporean adults than younger ones, such that older adults were more able to adapt their daily activities to their satisfaction^37, 38^. This, coupled with the emotional resilience and increased tendency to engage in proactive coping that older adults exhibit ^39–41^, may have enabled them to recover faster from symptoms of anxiety and depression during pandemic than younger adults^42^. The findings are also consistent with research regarding the natural lifespan of anxiety and depressive disorders, which find that symptoms of anxiety may be less common among older age groups as the prevalence of anxiety disorders tends to decrease with age^43^. However, literature suggests that older adults may be less emotionally but more physiologically reactive to stressors, such that they exhibit greater increases in their ambulatory blood pressure than younger adults in the face of stressors despite reporting a weaker impact of these stressors on their mood^44^. As such, concerns regarding the impact of COVID-19 psychological stressors on the well-being of older adults may not be unfounded, but more resources should be directed to younger adults in Singapore as a more psychologically vulnerable population in the face of the pandemic.

There are limitations of this study. Firstly, there was a lack of pre-COVID mental health data collected, and the absence of a pre-COVID baseline for depression and anxiety symptoms made it difficult to gauge the initial impact of COVID-19 on the population and therefore Singaporeans’ resilience to these symptoms in the face of the pandemic. However, analyses of mental health in Singapore following the Circuit Breaker are still informative given the differential trends in symptoms among younger and older adults. Secondly, the study data were collected online, such that data from older adults may be biased towards those who were more technologically savvy and therefore less impacted by the effects of isolation during the pandemic. Third, the PHQ-4 is a screening tool for depression and anxiety, rather than a diagnostic one, and therefore only reflects changes in the population’s symptoms rather than the prevalence of the disorders themselves. Furthermore, as there are only four questions in the questionnaire and each is only scored from zero to three, the scale may be relatively insensitive to changes in anxiety and depression symptoms. It remains a validated, widely accepted scale that suffices for the intent of the study, but further research is needed for trends in psychological symptoms using a more sensitive scale (such as the PHQ-9 or the GAD-7) and trends of actual mental diagnoses throughout the pandemic. Such research should also be conducted across a longer time period, as 17 months would not suffice to observe trends in chronic disorders rather than simply affective symptoms.

This study is the first to date to analyze trends in depression and anxiety symptoms by age groups in Singapore across Singapore’s pandemic restriction periods. Its findings could be used to inform future pandemic policies and to identify areas of psychological vulnerability in Singapore’s population during future epidemics and crises. In particular, these results suggest that Singapore’s younger adults may be particularly susceptible to worsening depressive and anxiety symptoms as the health crisis persists, which may exacerbate existing inequalities in mental health as younger adults already report more symptoms than older adults at the baseline (during Circuit Breaker; Supplementary Appendix Table S10) ^43^. The study thus highlights that more resources should be allocated to younger adults in the form of outreach or increased telehealth services should another pandemic occur.

## Supporting information

STROBE Checklist

## Data Availability

All data files and code required to reproduce these analyses are publicly available on the Imperial College GitHub site (https://github.com/mrc-ide/SGP_covid19_mental_health).

https://github.com/YouGov-Data/covid-19-tracker

## Role of the funding source

C.M. acknowledges funding from the Medical Research Council (MRC) Centre for Global Infectious Disease Analysis (MR/X020258/1) funded by the UK MRC and carried out in the frame of the Global Health EDCTP3 Joint Undertaking supported by the EU; a philanthropic donation from Community Jameel supporting the work of the Jameel Institute; the Schmidt Foundation (grant code 6–22–63345). S.H.X.T. is supported by the National University Health System Clinician Scientist Program (NCSP 2.0) AND National Medical Research Council, Singapore Research Training Fellowship Award. The funders of the study had no role in study design, data collection, data analysis, data interpretation, or writing of the report. For the purpose of open access, the author has applied a ‘Creative Commons Attribution’ (CC BY) licence to any Author Accepted Manuscript version arising from this submission.

Since the ICL-YouGov data used in the study was publicly available, collected prior to the study, and not individually traceable, the study was exempt from ethics review.

## Contributions

S.H.X.T., C.M., K.B.T., N.S.E.G., and A.J.W.Y. conceptualised the study, and were involved in methodology and reviewing and editing the manuscript. S.H.X.T., C.M., and K.B.T. were responsible for project administration and study supervision. N.S.E.G. was responsible for data curation and visualisation, formal analysis, and writing of the original draft. A.J.W.Y. contributed to data verification, curation, and validation. C.M. was responsible for survey resources and contributed to data collection. S.C.Y.W., M.S., E.B.A., and D.C. B.L. interpreted the data, and reviewed and edited the manuscript. All authors contributed comments to the paper, had access to the raw data, and agree to be accountable for all aspects of the work. The corresponding author had final responsibility to submit for publication.

## References

1. National Institutes of Health (NIH) [Internet]. [cited 2024 Jun 14]. National Institutes of Health (NIH). Available from: https://www.nih.gov/

2. Fountoulakis KN, Apostolidou MK, Atsiova MB, Filippidou AK, Florou AK, Gousiou DS, et al. Self-reported changes in anxiety, depression and suicidality during the COVID-19 lockdown in Greece. Journal of Affective Disorders. 2021 Jan 15;279:624–9.

3. Bignardi G, Dalmaijer ES, Anwyl-Irvine AL, Smith TA, Siugzdaite R, Uh S, et al. Longitudinal increases in childhood depression symptoms during the COVID-19 lockdown. Arch Dis Child. 2020 Dec 9;106(8):791–7.

4. Daly M, Robinson E. Psychological distress and adaptation to the COVID-19 crisis in the United States. Journal of Psychiatric Research. 2021 Apr 1;136:603–9.

5. Liang L, Ren H, Cao R, Hu Y, Qin Z, Li C, et al. The Effect of COVID-19 on Youth Mental Health. Psychiatr Q. 2020;91(3):841–52.

6. Faris M, Macky MM, Badran AH, Saif M, Yasser M, Ibrahim E, et al. The Prevalence of Anxiety Among University Students in the United Arab Emirates Following the COVID-19 Lockdown. Cureus. 2024 Mar;16(3):e56259.

7. Cao W, Fang Z, Hou G, Han M, Xu X, Dong J, et al. The psychological impact of the COVID-19 epidemic on college students in China. Psychiatry Research. 2020 May 1;287:112934.

8. Ahorsu DK, Lin CY, Imani V, Saffari M, Griffiths MD, Pakpour AH. The Fear of COVID-19 Scale: Development and Initial Validation. Int J Ment Health Addiction. 2022 Jun 1;20(3):1537–45.

9. Aknin LB, Andretti B, Goldszmidt R, Helliwell JF, Petherick A, De Neve JE, Dunn EW, Fancourt D, Goldberg E, Jones SP, Karadag O. Policy stringency and mental health during the COVID-19 pandemic: a longitudinal analysis of data from 15 countries. The Lancet Public Health. 2022 May 1;7(5):e417–26.

10. Lei L, Huang X, Zhang S, Yang J, Yang L, Xu M. Comparison of Prevalence and Associated Factors of Anxiety and Depression Among People Affected by versus People Unaffected by Quarantine During the COVID-19 Epidemic in Southwestern China. Med Sci Monit. 2020 Apr 26;26:e924609.

11. Saltzman LY, Lesen AE, Henry V, Hansel TC, Bordnick PS. COVID-19 Mental Health Disparities. Health Security. 2021 Jun;19(S1):S–5.

12. Achterberg M, Dobbelaar S, Boer OD, Crone EA. Perceived stress as mediator for longitudinal effects of the COVID-19 lockdown on wellbeing of parents and children. Sci Rep. 2021 Feb 3;11(1):2971.

13. Francisco R, Pedro M, Delvecchio E, Espada JP, Morales A, Mazzeschi C, et al. Psychological Symptoms and Behavioral Changes in Children and Adolescents During the Early Phase of COVID-19 Quarantine in Three European Countries. Front Psychiatry. 2020 Dec 3;11:570164.

14. Pedersen MT, Andersen TO, Clotworthy A, Jensen AK, Strandberg-Larsen K, Rod NH, et al. Time trends in mental health indicators during the initial 16 months of the COVID-19 pandemic in Denmark. BMC Psychiatry. 2022 Jan 10;22(1):25.

15. Oliu-Barton M, Pradelski BS, Aghion P, Artus P, Kickbusch I, Lazarus JV, Sridhar D, Vanderslott S. SARS-CoV-2 elimination, not mitigation, creates best outcomes for health, the economy, and civil liberties. The Lancet. 2021 Jun 12;397(10291):2234–6.

16. Amicucci G, Salfi F, D’Atri A, Viselli L, Ferrara M. The Differential Impact of COVID-19 Lockdown on Sleep Quality, Insomnia, Depression, Stress, and Anxiety among Late Adolescents and Elderly in Italy. Brain Sci. 2021 Oct 11;11(10):1336.

17. Klaiber P, Wen JH, DeLongis A, Sin NL. The Ups and Downs of Daily Life During COVID-19: Age Differences in Affect, Stress, and Positive Events. J Gerontol B Psychol Sci Soc Sci. 2021 Jan 18;76(2):e30–7.

18. Burr DA, Castrellon JJ, Zald DH, Samanez-Larkin GR. Emotion dynamics across adulthood in everyday life: Older adults are more emotionally stable and better at regulating desires. Emotion. 2021 Apr;21(3):453–64.

19. Carstensen LL, Pasupathi M, Mayr U, Nesselroade JR. Emotional experience in everyday life across the adult life span. J Pers Soc Psychol. 2000 Oct;79(4):644–55.

20. Janiri D, Kotzalidis GD, Giuseppin G, Molinaro M, Modica M, Montanari S, et al. Psychological Distress After Covid-19 Recovery: Reciprocal Effects With Temperament and Emotional Dysregulation. An Exploratory Study of Patients Over 60 Years of Age Assessed in a Post-acute Care Service. Front Psychiatry [Internet]. 2020 Nov 12 [cited 2024 Jun 14];11. Available from: https://www.frontiersin.org/journals/psychiatry/articles/10.3389/fpsyt.2020.590135/full

21. YouGov-Data/covid-19-tracker [Internet]. YouGov Data; 2024 [cited 2024 Jun 21]. Available from: https://github.com/YouGov-Data/covid-19-tracker

22. Sung SC, Low CCH, Fung DSS, Chan YH. Screening for major and minor depression in a multiethnic sample of Asian primary care patients: a comparison of the nine-item Patient Health Questionnaire (PHQ-9) and the 16-item Quick Inventory of Depressive Symptomatology - Self-Report (QIDS-SR16). Asia Pac Psychiatry. 2013 Dec;5(4):249–58.

23. Kroenke K, Spitzer RL, Williams JBW, Löwe B. An Ultra-Brief Screening Scale for Anxiety and Depression: The PHQ–4. Psychosomatics. 2009 Nov 1;50(6):613–21.

24. Levis B, Sun Y, He C, Wu Y, Krishnan A, Bhandari PM, et al. Accuracy of the PHQ-2 Alone and in Combination With the PHQ-9 for Screening to Detect Major Depression: Systematic Review and Meta-analysis. JAMA. 2020 Jun 9;323(22):2290–300.

25. Wilson JM, Lee J, Shook NJ. COVID-19 worries and mental health: the moderating effect of age. Aging & Mental Health. 2021 Jul 3;25(7):1289–96.

26. Fairlie R. The impact of COVID□19 on small business owners: Evidence from the first three□months after widespread social□distancing restrictions. J Econ Manag Strategy. 2020;29(4):727–40.

27. Dunphy C, Miller GF, Rice K, Vo L, Sunshine G, McCord R, et al. The Impact of Covid-19 State Closure Orders on Consumer Spending, Employment, and Business Revenue. Journal of Public Health Management and Practice. 2022 Feb;28(1):43.

28. Imran A. Evaluating Wellbeing and Worries of University Students during COVID-19 Pandemic [Internet]. [cited 2024 Jun 14]. Available from: https://dergipark.org.tr/en/pub/atauniiibd/issue/59801/776979

29. Wathelet M, Duhem S, Vaiva G, Baubet T, Habran E, Veerapa E, et al. Factors Associated With Mental Health Disorders Among University Students in France Confined During the COVID-19 Pandemic. JAMA Netw Open. 2020 Oct 1;3(10):e2025591.

30. Fruehwirth JC, Biswas S, Perreira KM. The Covid-19 pandemic and mental health of first-year college students: Examining the effect of Covid-19 stressors using longitudinal data. PLOS ONE. 2021 Mar 5;16(3):e0247999.

31. Choi H, Irwin MR, Cho HJ. Impact of social isolation on behavioral health in elderly: Systematic review. World J Psychiatry. 2015 Dec 22;5(4):432–8.

32. Beam CR, Kim AJ. Psychological sequelae of social isolation and loneliness might be a larger problem in young adults than older adults. Psychol Trauma. 2020 Aug;12(S1):S58–60.

33. Nyqvist F, Victor CR, Forsman AK, Cattan M. The association between social capital and loneliness in different age groups: a population-based study in Western Finland. BMC Public Health. 2016 Jul 11;16(1):542.

34. Luchetti M, Lee JH, Aschwanden D, Sesker A, Strickhouser JE, Terracciano A, et al. The Trajectory of Loneliness in Response to COVID-19. Am Psychol. 2020 Oct;75(7):897–908.

35. Hämmig O. Health risks associated with social isolation in general and in young, middle and old age. PLoS One. 2019 Jul 18;14(7):e0219663.

36. Loades ME, Chatburn E, Higson-Sweeney N, Reynolds S, Shafran R, Brigden A, et al. Rapid Systematic Review: The Impact of Social Isolation and Loneliness on the Mental Health of Children and Adolescents in the Context of COVID-19. J Am Acad Child Adolesc Psychiatry. 2020 Nov;59(11):1218–1239.e3.

37. Yang HC, Thornton LM, Shapiro CL, Andersen BL. Surviving recurrence: Psychological and quality-of-life recovery. Cancer. 2008;112(5):1178–87.

38. Yu CC, Tou NX, Low JA. A comparative study on mental health and adaptability between older and younger adults during the COVID-19 circuit breaker in Singapore. BMC Public Health. 2022 Mar 15;22(1):507.

39. Carstensen LL, Shavit YZ, Barnes JT. Age Advantages in Emotional Experience Persist Even Under Threat From the COVID-19 Pandemic. Psychol Sci. 2020 Nov;31(11):1374–85.

40. Pierce M, Hope H, Ford T, Hatch S, Hotopf M, John A, et al. Mental health before and during the COVID-19 pandemic: a longitudinal probability sample survey of the UK population. The Lancet Psychiatry. 2020 Oct 1;7(10):883–92.

41. Pieh C, Budimir S, Delgadillo J, Barkham M, Fontaine JRJ, Probst T. Mental Health During COVID-19 Lockdown in the United Kingdom. Psychosomatic Medicine. 2021 May;83(4):328.

42. Pearman A, Hughes ML, Smith EL, Neupert SD. Age Differences in Risk and Resilience Factors in COVID-19-Related Stress. J Gerontol B Psychol Sci Soc Sci. 2021 Jan 18;76(2):e38–44.

43. Bandelow B, Michaelis S. Epidemiology of anxiety disorders in the 21st century. Dialogues in clinical neuroscience. 2015 Sep 30;17(3):327–35.

44. Uchino BN, Berg CA, Smith TW, Pearce G, Skinner M. Age-related differences in ambulatory blood pressure during daily stress: Evidence for greater blood pressure reactivity with age. Psychology and Aging. 2006;21(2):231–9.

