## Supplementary material for "Inequalities in Mental Health: Age-related Trends Across Pandemic Phases in Singapore": Figures and Tables

**Figure 1. Pandemic measure breakdown for the study duration in Singapore.**

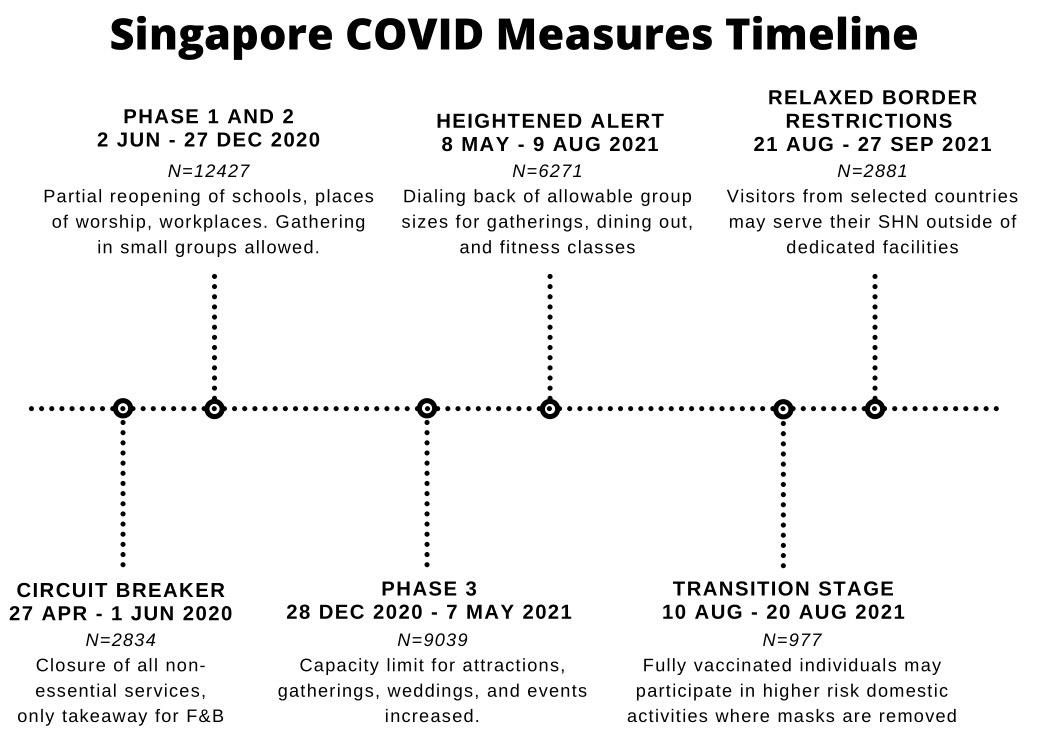

Figure 1. Pandemic measure breakdown for the study duration in Singapore. Participants were in Singapore and above the age of 18. Restriction periods were grouped based on restriction types and types of non-pharmaceutical interventions.

**Table 1.** **Demographic characteristics of survey population across 36 waves of the COVID-19 behavioral tracker in Singapore.**

|  | **Sample characteristics** | **PHQ-4 total score** | **GAD-2** | **PHQ-2** |
| --- | --- | --- | --- | --- |
|  | **Count (%)** | **Mean (SD)** | **Percentage with Significant Symptoms (SD)** | |
| *Full Population* |  |  |  |  |
|  | 34429 (100%) | 3·51 (3·27) | 0·26 (0·44) | 0·26 (0·44) |
| *Age group* |  |  |  |  |
| 18–29 | 6999 (20·33%) | 4·71 (3·17) | 0·38 (0·49) | 0·37 (0·48) |
| 30–39 | 7401 (21·50%) | 4·16 (3·21) | 0·32 (0·47) | 0·32 (0·47) |
| 40–49 | 7006 (20·35%) | 3·54 (3·22) | 0·25 (0·43) | 0·26 (0·44) |
| 50–59 | 7099 (20·62%) | 2·83 (3·16) | 0·20 (0·40) | 0·19 (0·39) |
| 60+ | 5924 (17·21%) | 2·06 (2·90) | 0·14 (0·35) | 0·14 (0·35) |
| *Sex* |  |  |  |  |
| Female | 17176 (49·89%) | 3·52 (3·21) | 0·26 (0·44) | 0·24 (0·43) |
| Male | 17253 (50·11%) | 3·50 (3·33) | 0·26 (0·44) | 0·27 (0·45) |
| *Have mental health condition* | | |  |  |
| No | 28996 (84·22%) | 3·37 (3·23) | 0·25 (0·43) | 0·25 (0·43) |
| Prefer not to say | 4857 (14·11%) | 4·00 (3·28) | 0·31 (0·46) | 0·30 (0·46) |
| Yes | 576 (1·67%) | 6·06 (3·77) | 0·52 (0·50) | 0·53 (0·50) |
| *Employment status* | | |  |  |
| Full time employment | 21990 (63·87%) | 3·60 (3·20) | 0·27 (0·44) | 0·26 (0·44) |
| Not working | 3865 (11·23%) | 2·19 (2·97) | 0·15 (0·36) | 0·15 (0·36) |
| Other | 6572 (19·09%) | 3·66 (3·31) | 0·28 (0·45) | 0·27 (0·45) |
| Unemployed | 2002 (5·81%) | 4·57 (3·72) | 0·35 (0·48) | 0·37 (0·48) |

Table 1. Demographic characteristics of survey population across 36 waves of the COVID-19 behavioral tracker in Singapore. Younger adults experienced more symptoms of depression and anxiety than older adults. Those with mental health conditions and those who were unemployed reported levels of symptoms that were higher than the population average.

**Table 2. Multiple linear regression of the change in psychological distress (total PHQ-4 score) from Circuit Breaker to each restriction period for each age group.**

|  | Coefficients (95% CI) | | | | | |
| --- | --- | --- | --- | --- | --- | --- |
|  | Change in Anxiety and Depression Symptoms (PHQ-4) for Ages: | | | | | |
|  | ***Full Population*** | **Ages 18 to 29** | **Ages 30 to 39** | **Ages 40 to 49** | **Ages 50 to 59** | **Ages Over 60** |
| **Circuit**  **Breaker** | *Reference* | *Reference* | *Reference* | *Reference* | *Reference* | *Reference* |
| **Phase**  **1 and 2** | *-0·06 (-0·19, 0·06)* | 0·23 (-0·05, 0·52) | 0·15 (-0·13, 0·43) | -0·11 (-0·39, 0·18) | -0·33 (-0·60, -0·06) * | -0·23 (-0·52, 0·05) |
| **Phase 3** | *-0·10 (-0·24, 0·03)* | 0·32 (-0·00, 0·65) | 0·40 (0·08, 0·71) * | -0·08 (-0·40, 0·25) | -0·54 (-0·85, -0·22) * | -0·39 (-0·72, -0·07) * |
| **Heightened**  **Alert** | *-0·05 (-0·19, 0·09)* | 0·40 (0·08, 0·73) * | 0·29 (-0·02, 0·60) | -0·06 (-0·38, 0·26) | -0·32 (-0·61, -0·02) * | -0·44 (-0·75, -0·14) * |
| **Transition** | *-0·08 (-0·31, 0·14)* | 0·27 (-0·25, 0·79) | 0·20 (-0·30, 0·69) | -0·07 (-0·58, 0·44) | -0·21 (-0·71, 0·30) | -0·53 (-1·03, -0·04) * |
| **Relaxed**  **Border**  **Restrictions** | *0·00 (-0·16, 0·16)* | 0·59 (0·22, 0·97) * | 0·44 (0·08, 0·80) * | -0·09 (-0·46, 0·29) | -0·45 (-0·80, -0·10) * | -0·38 (-0·73, -0·02) * |

Table 2. Multiple linear regression of the change in psychological distress (total PHQ-4 score) from Circuit Breaker to each restriction period for each age group. All models are adjusted for covariates (age, sex, mental health status, employment status, and availability of COVID-19 vaccine). As restrictions were lifted, younger adults tended to report more anxiety and depression symptoms, but older adults tended to report fewer.

**P < 0·05*

**Table 3. Multiple logistic regression of the change in symptoms of anxiety (3 or higher on the GAD-2) from Circuit Breaker to each restriction period for each age group.**

|  | Odds Ratio (95% CI) | | | | | |
| --- | --- | --- | --- | --- | --- | --- |
|  | Change in Anxiety Symptoms (GAD-2) for Ages: | | | | | |
|  | ***Full Population*** | **Ages 18 to 29** | **Ages 30 to 39** | **Ages 40 to 49** | **Ages 50 to 59** | **Ages Over 60** |
| **Circuit**  **Breaker** | *Reference* | *Reference* | *Reference* | *Reference* | *Reference* | *Reference* |
| **Phase**  **1 and 2** | 0·98 (0·89, 1·08) | 1·18 (0·97, 1·44) | 1·23 (1·01, 1·50) * | 0·99 (0·80, 1·23) | 0·75 (0·61, 0·92) * | 0·71 (0·56, 0·92) * |
| **Phase 3** | 1·00 (0·90, 1·12) | 1·28 (1·02, 1·60) * | 1·41 (1·12, 1·76) * | 0·95 (0·75, 1·21) | 0·68 (0·53, 0·88) * | 0·69 (0·51, 0·92) * |
| **Heightened**  **Alert** | 0·97 (0·88, 1·08) | 1·25 (1·01, 1·56) * | 1·22 (0·98, 1·53) | 1·00 (0·79, 1·27) | 0·77 (0·61, 0·97) * | 0·62 (0·47, 0·81) * |
| **Transition** | 0·92 (0·78, 1·10) | 1·28 (0·90, 1·81) | 1·01 (0·71, 1·44) | 1·02 (0·70, 1·49) | 0·86 (0·58, 1·27) | 0·44 (0·26, 0·75) * |
| **Relaxed**  **Border**  **Restrictions** | 1·03 (0·91, 1·16) | 1·48 (1·15, 1·91) * | 1·29 (1·00, 1·66) | 1·03 (0·78, 1·36) | 0·79 (0·60, 1·03) * | 0·60 (0·43, 0·83) * |

Table 3. Multiple logistic regression of the change in symptoms of anxiety (3 or higher on the GAD-2) from Circuit Breaker to each restriction period for each age group. All models are adjusted for covariates (age, sex, mental health status, employment status, and availability of COVID-19 vaccine). Younger adults were more likely to report symptoms of anxiety when restrictions were lifted, but older adults were less likely to do so.

**P < 0·05*

**Table 4. Multiple logistic regression of the change in symptoms of depression (3 or higher on the PHQ2) from Circuit Breaker to each restriction period for each age group.**

|  | Odds Ratio (95% CI) | | | | | | | | | | |
| --- | --- | --- | --- | --- | --- | --- | --- | --- | --- | --- | --- |
|  | Change in Depression Symptoms (PHQ-2) for Ages: | | | | | | | | | | |
|  | ***Full Population*** | | **Ages 18 to 29** | | **Ages 30 to 39** | | **Ages 40 to 49** | | **Ages 50 to 59** | | **Ages Over 60** |
| **Circuit**  **Breaker** | *Reference* | *Reference* | | *Reference* | | *Reference* | | *Reference* | | *Reference* | |
| **Phase**  **1 and 2** | 0·87 (0·79, 0·96) * | 0·94 (0·77, 1·14) | | 0·90 (0·74, 1·10) | | 0·86 (0·70, 1·06) | | 0·75 (0·61, 0·93) * | | 0·90 (0·69, 1·16) | |
| **Phase 3** | 0·87 (0·78, 0·97) * | 1·02 (0·82, 1·27) | | 1·01 (0·81, 1·26) | | 0·91 (0·72, 1·16) | | 0·63 (0·49, 0·81) * | | 0·73 (0·54, 0·98) * | |
| **Heightened**  **Alert** | 0·85 (0·77, 0·94) * | 1·01 (0·81, 1·26) | | 0·89 (0·71, 1·10) | | 0·86 (0·68, 1·08) | | 0·78 (0·62, 0·98) * | | 0·69 (0·52, 0·92) * | |
| **Transition** | 0·80 (0·68, 0·96) * | 0·93 (0·66, 1·33) | | 0·92 (0·65, 1·30) | | 0·79 (0·54, 1·17) | | 0·82 (0·55, 1·22) | | 0·48 (0·28, 0·83) * | |
| **Relaxed**  **Border**  **Restrictions** | 0·87 (0·77, 0·98) * | 1·13 (0·87, 1·45) | | 1·05 (0·82, 1·34) | | 0·86 (0·65, 1·13) | | 0·64 (0·48, 0·85) * | | 0·64 (0·45, 0·91) * | |

Table 4. Multiple logistic regression of the change in symptoms of depression (3 or higher on the PHQ2) from Circuit Breaker to each restriction period for each age group. All models are adjusted for covariates (age, sex, mental health status, employment status, and availability of COVID-19 vaccine). Older adults were less likely to report depressive symptoms later in the pandemic.

**P < 0·05*
