## Supplementary material for "Inequalities in Mental Health: Age-related Trends Across Pandemic Phases in Singapore": STROBE Checklist

**STROBE Statement**

|  | Item No. | Recommendation | | Relevant text from manuscript |
| --- | --- | --- | --- | --- |
| **Title and abstract** | 1 | (*a*) Indicate the study’s design with a commonly used term in the title or the abstract | | Abstract: Methods, Page 2, Lines 8–11 |
|  |  | (*b*) Provide in the abstract an informative and balanced summary of what was done and what was found | | Abstract: Methods, Page 2, Lines 6–11  Abstract: Findings, Page 2, Lines 13–18  Abstract: Interpretation, Page 2, Lines 20–22 |
| Background/rationale | 2 | Explain the scientific background and rationale for the investigation being reported | | Research in context: Evidence before this study, Page 3–4  Introduction, Page 3, Lines 27–68 |
| Objectives | 3 | State specific objectives, including any prespecified hypotheses | | Introduction, Page 3, Lines 61–68 |
| Study design | 4 | Present key elements of study design early in the paper | | Methods and Measures, Page 4–5, Lines 70–122 |
| Setting | 5 | Describe the setting, locations, and relevant dates, including periods of recruitment, exposure, follow-up, and data collection | | Methods and Measures, Page 4, Lines 71–77 |
| Participants | 6 | Give the eligibility criteria, and the sources and methods of selection of participants | | Methods and Measures, Page 4, Lines 71–77  Supplementary Appendix 1.1. Data, detailed information, Page 2–3 |
| Variables | 7 | Clearly define all outcomes, exposures, predictors, potential confounders, and effect modifiers. Give diagnostic criteria, if applicable | | Methods and Measures, Page 4–5, Lines 79–122 |
| Data sources/ measurement | 8 | For each variable of interest, give sources of data and details of methods of assessment (measurement). Describe comparability of assessment methods if there is more than one group | | Methods and Measures, Page 4–5, Lines 79–122  Supplementary Appendix: 1.1. Data, detailed information, Page 2 |
| Bias | 9 | Describe any efforts to address potential sources of bias | | Methods and Measures, Page 5, Lines 116–122 |
| Study size | 10 | Explain how the study size was arrived at | | Methods and Measures, Page 4, Lines 74–77  Supplementary Appendix: 1.1. Data, detailed information |

Continued on next page

| Quantitative variables | 11 | Explain how quantitative variables were handled in the analyses. If applicable, describe which groupings were chosen and why | Methods and Measures, Page 5, Lines 107–122  Supplementary Appendix: 1.1. Data, detailed information, Page 2 |
| --- | --- | --- | --- |
| Statistical methods | 12 | (*a*) Describe all statistical methods, including those used to control for confounding | Methods and Measures, Page 5, Lines 107–122  Supplementary Appendix: 1.2. Model selection, Page 6–9 |
|  |  | (*b*) Describe any methods used to examine subgroups and interactions | Methods and Measures, Page 5, Lines 116–122  Supplementary Appendix: 1.2. Model selection, Page 6–9 |
|  |  | (*c*) Explain how missing data were addressed | Methods and Measures, Page 4, Lines 74–77  Supplementary Appendix: 1.1. Data, detailed information, Page 2–3 |
|  |  | (*d*) If applicable, describe analytical methods taking account of sampling strategy | Methods and Measures, Page 5, Lines 116–122 |
|  |  | (*e*) Describe any sensitivity analyses | Supplementary Appendix: 2. Results, Page 6–9 |
| Participants | 13 | (a) Report numbers of individuals at each stage of study—eg numbers potentially eligible, examined for eligibility, confirmed eligible, included in the study, completing follow-up, and analysed | Methods and Measures, Page 4, Lines 71–77  Supplementary Appendix: 1.1. Data, detailed information, Page 1–2 |
|  |  | (b) Give reasons for non-participation at each stage | Methods and Measures Page 4–5, Lines 74–77, 107–115  Supplementary Appendix: 1.1. Data, detailed information, Page 1–2 |
|  |  | (c) Consider use of a flow diagram | Supplementary Appendix: 1.1. Data, detailed information, Page 2 |
| Descriptive data | 14 | (a) Give characteristics of study participants (eg demographic, clinical, social) and information on exposures and potential confounders | Results, Page 5, Lines 124–130  Supplementary Appendix: 1.2 Demographic breakdowns, Page 10–14 |
|  |  | (b) Indicate number of participants with missing data for each variable of interest | Methods and Measures, Page 4–5, Lines 71–77, 107–115  Supplementary Appendix: 1.1. Data, detailed information, Page 2–3 |
| Outcome data | 15 | Report numbers of outcome events or summary measures | Results, Page 5–6, Lines 124–171 |
| Main results | 16 | (*a*) Give unadjusted estimates and, if applicable, confounder-adjusted estimates and their precision (eg, 95% confidence interval). Make clear which confounders were adjusted for and why they were included | Methods and Measures, Page 5, Lines 116–122  Results, Page 5–6, Lines 121–171 |
|  |  | (*b*) Report category boundaries when continuous variables were categorized | Methods and Measures, Page 5, Lines 107–115  Results, Page 5–6, Lines 121–171 |
|  |  | (*c*) If relevant, consider translating estimates of relative risk into absolute risk for a meaningful time period | NA |

Continued on next page

| Other analyses | 17 | Report other analyses done—eg analyses of subgroups and interactions, and sensitivity analyses | Results, Pages 5–6, Lines 145–151  Supplementary Appendix: 1.3 Model Selection, Page 6–9  Supplementary Appendix: 2. Results, Page 12–17 |
| --- | --- | --- | --- |
| Key results | 18 | Summarise key results with reference to study objectives | Discussion, Page 6–7, Lines 173–217 |
| Limitations | 19 | Discuss limitations of the study, taking into account sources of potential bias or imprecision. Discuss both direction and magnitude of any potential bias | Discussion, Page 7, Lines 218–232 |
| Interpretation | 20 | Give a cautious overall interpretation of results considering objectives, limitations, multiplicity of analyses, results from similar studies, and other relevant evidence | Discussion, Page 6–7, Lines 173–241 |
| Generalisability | 21 | Discuss the generalisability (external validity) of the study results | Discussion, Page 6–7, Lines 173–180, 233–241 |
| Other information | |  |  |
| Funding | 22 | Give the source of funding and the role of the funders for the present study and, if applicable, for the original study on which the present article is based | Role of the Funding Source, Page 8, Lines 247–256 |

**Note:** An Explanation and Elaboration article discusses each checklist item and gives methodological background and published examples of transparent reporting. The STROBE checklist is best used in conjunction with this article (freely available on the Web sites of PLoS Medicine at http://www.plosmedicine.org/, Annals of Internal Medicine at http://www.annals.org/, and Epidemiology at http://www.epidem.com/). Information on the STROBE Initiative is available at www.strobe-statement.org.
