## Supplementary Appendix for "Inequalities in Mental Health: Age-related Trends Across Pandemic Phases in Singapore"

| 1. Methods | 2 |
| --- | --- |
| 1.1 Data, detailed information | 2 |
| 1.2 Restriction Periods | 4 |
| 1.3 Model selection | 6 |
| 2. Results | 10 |
| 2.1 Demographic breakdowns | 10 |
| 2.2 Additional regressions | 14 |
| 2.3 Baseline Comparisons | 16 |

**Supplementary Appendix**

**1. Methods**

**1.1 Data, detailed information**

In each of the latter 36 waves of the YouGov survey, participants were asked to fill out the PHQ-4 questionnaire. The exact questions are as follows: Over the last two weeks, how often have you been bothered by the following problems? 1. Feeling nervous, anxious or on edge. 2. Not being able to stop or control worrying 3. Feeling down, depressed or hopeless. And 4. Little interest or pleasure in doing things. The first two questions make up the GAD-2, while the latter two make up the PHQ-2. Scores in the PHQ-4 were summed up to create a numeric PHQ-4 Total Score variable that ranged from 0 to 12. Scores in the PHQ-2 and GAD-2 were summed up and coded as “1” for significant symptoms if they were greater than or equal to 3, and “0” otherwise.

The YouGov survey collected demographic data on participants’ genders, ages, employment status, as well as whether the participant had ever been diagnosed with a chronic condition (“Which, if any, of the following have you been diagnosed with?” Please select all that apply: Arthritis / Asthma / Cancer / Cystic fibrosis / Chronic obstructive pulmonary disease (COPD) / Diabetes / Epilepsy / Heart disease / High blood pressure / High cholesterol / HIV/ Aids / Mental health condition / Multiple Sclerosis / Prefer not to say / None of these). A new variable, Mental Health Condition, was created and used in the analysis, with “Yes” indicating that a person had previously been diagnosed with a mental health condition, “No” indicating that they had not, and “Prefer not to say” indicating that no response was given to the survey question.

Data was also collected on whether vaccinations were available to the participant (“COVID-19 vaccination is not yet available for me.” No / Yes). This variable was reverse coded such that “Yes” indicated that vaccination was available to the respondent and “No” indicated that it was not.

No other transformations were applied to the data outside of the category groupings described in the paper’s methods section. Data was cleaned according to the flowchart below (Figure F1).

**Figure S1. Study Flowchart**

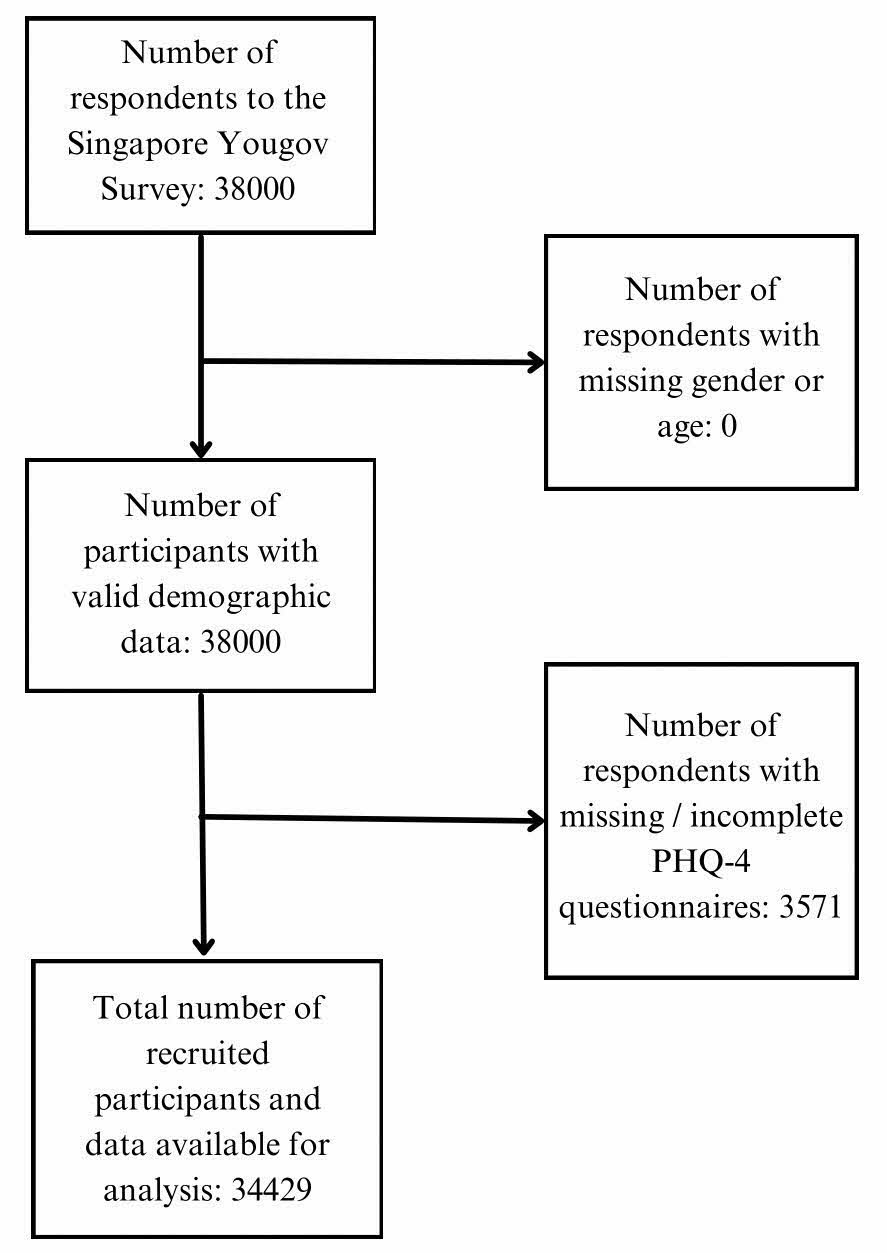

**1.2 Restriction Periods**

**Table S5. Detailed description of restriction measures and mean Oxford COVID-19 Government Response Tracker (OxCGRT) Stringency Index scores for each measure period**

| **Date** | **Measure Period** | **Restrictions Implemented** | **OxCGRT Stringency Index Average** |
| --- | --- | --- | --- |
| 03 Apr 2020 | Circuit Breaker | Implementation of Circuit Breaker (CB) measures from 07 Apr 2020 onwards. Measures include closures of workplaces, schools, recreation venues, places of worship. Only essential services were allowed, with only option of takeaway/delivery for food and beverage (F&B) | 79.88998 |
| 2 Jun 2020 | Phase 1 [Safe-Reopening] | 1. More Singaporeans can go to work 2. Businesses stagger working hours to minimize travel during peak hours  3. Marriage solemnizations - up to 10 people are permitted 4. Places of worship for private worship, up to 5 members from same household 5. Preschools gradually open 6. Primary and secondary graduating cohorts attend school daily, other cohorts alternate weekly 7. Junior Colleges and Millenia Institute: half students back at any point, institutes of higher learning will have lectures online, co-curricular activities will not resume 8. Allowed to visit other family households (grandparents, etc.) subject to two visitors from same household per day 9. Important hospital-based care and dental care will gradually resume on a case-by-case basis. | 55.49588 |
| 19 Jun 2020 | Phase 2 | 1. Gatherings of 5 people may resume 2. Retail businesses may open physical outlets 3. F&B dine-in can resume, alcohol ceasing at 2230HRS 4. Live music, television and video screenings not allowed 5. Sports, parks, other public facilities including swimming pools will be open |  |
| 28 Dec 2020 | Phase 3 | 1. Social gathering limit increased to 8 persons, from 5 2. Capacity limit increased to 8 square meters/person, from 10, attractions up to 65% operating capacity, from 50% 3. ROs capacity increased to 250, with allowance of live performance elements 4. Marriage solemnizations visitor limit increased to 10 persons, from 8 5. Audience limit for live performances in Arts and Culture sector increased to 250, from 100 | 51.02087 |
| 8 May 2021 | Phase 3 Heightened Alert | 1. Group sizes reduced from 8 pax to 5 pax. Maximum 5 unique household visitors a day. 2. No more than 50% of the workforce who may work at the workplace (down from 75%). 3. Congregational and worship services suspend masked congregational singing. Maximum 250 pax gatherings with pre-event test (PET), max 100 pax without PET. 4, Capacity of museums, public libraries, attractions and shows reduced operating capacity to 50% (from 65%). Maximum of 100 pax without PET for shows and attractions. 5. Indoor gymnasiums and fitness studios closed to public. All mass participation sports events suspended. | 54.93269 |
| 19 July 2021 | Phase 3 Heightened Alert (Changes) | 1. Dine-in dialed back to groups of 2 persons, but vaccinated individuals, unvaccinated individuals with a valid negative pre-event test result, children aged 12 years and below or recovered individuals can dine-in in groups of up to 5 persons. 2. Group size for mask-off fitness classes reduced to 2 people per group (down from 5 a group) |  |
| 22 July 2021 | Phase 2 Heightened Alert | 1. Group sizes for social gatherings reduced from maximum of 5 persons to maximum of 2 persons. 2. Dine-in at F&B establishments and mask-off activities will not be allowed. 3. Weddings can go on with maximum 100 pax with PET. |  |
| 10 August 2021 | Transition Stage begins | 1. Increase in social gathering group size limit from 2 persons to 5 persons.  2. Fully vaccinated individuals may participate in higher-risk activities where masks are removed (e.g., dine-in) in a maximum of groups of 5. 3. Both vaccinated and unvaccinated people can dine-in at hawker centers and coffee shops in a maximum of groups of 2.  Individuals fully vaccinated with World Health Organization Emergency Use Listing Procedure vaccines such as Sinovac, Sinopharm and AstraZeneca will be considered "fully vaccinated" 2 weeks post receiving the full regimen of the vaccine.  Singapore citizens, permanent residents and long-term pass holders aged 12 years old and above who have not received a first dose of any COVID-19 vaccine can walk in to any of the 26 vaccination centers of Pfizer vaccine without prior appointment. | 44.46918 |
| 10 September 2021 | Transition Stage with changes to travel restrictions | Travelers from category 1 countries do not require SHN, category 2 countries can opt to serve their Stay-Home Notice (SHN) at home. | 43.90136 |

**1.3 Model selection**

The final prediction model was selected using the Akaike Information Criterion (AIC). Variables were added as shown in the table below (Table S1).

**Table S1. Model Selection Using AIC**

| **Variables Included in Model** | **Degrees of Freedom** | **AIC** |
| --- | --- | --- |
| Restriction Period | 7 | 179684.9 |
| Restriction Period, Gender | 8 | 179683.7 |
| Restriction Period, Gender, Age | 12 | 176728.6 |
| Restriction Period, Gender, Age, Employment Status | 15 | 176339.7 |
| Restriction Period, Gender, Age, Employment Status, Diagnosed with Mental Health Condition | 17 | 176006.1 |
| Restriction Period, Gender, Age, Employment Status, Diagnosed with Mental Health Condition, Vaccine Availability | 18 | 176006.3 |

The addition of vaccine availability as a variable slightly increased the model AIC from 176006.1 to 176006.3 (Table S1). The variable was retained in the model as the study was still concerned with how vaccine availability would impact mental health. Robustness checks were conducted, and the results of the best fit models are reported in tables S2 to S4. The best fit model measures the outcome variables against COVID restriction period accounting for gender, age group, employment status, and whether the participant was diagnosed with a mental health condition. The results are consistent with those reported in the paper.

**Table S2. Regression coefficients of the change in symptoms of anxiety and depression (total PHQ-4 score) from Circuit Breaker to each restriction period for each age group.**

|  | **Coefficients (95% CI)** | | | | | |
| --- | --- | --- | --- | --- | --- | --- |
|  | **Change in Anxiety and Depression Symptoms (PHQ-4) for Ages:** | | | | | |
|  | ***Full Population*** | **Ages 18 to 29** | **Ages 30 to 39** | **Ages 40 to 49** | **Ages 50 to 59** | **Ages Over 60** |
| **Circuit Breaker** | *Reference* | *Reference* | *Reference* | *Reference* | *Reference* | *Reference* |
| **Phase  1 and 2** | -0.06 (-0.19, 0.06) | 0.23 (-0.05, 0.52) | 0.15 (-0.13, 0.43) | -0.11 (-0.39, 0.18) | -0.33 (-0.60, -0.06) * | -0.23 (-0.52, 0.05) |
| **Phase 3** | -0.10 (-0.24, 0.03) | 0.35 (0.05, 0.64) * | 0.34 (0.04, 0.63) * | -0.16 (-0.46, 0.14) | -0.53 (-0.81, -0.24) * | -0.45 (-0.75, -0.16) * |
| **Heightened  alert** | -0.05 (-0.19, 0.09) | 0.40 (0.08, 0.73) * | 0.29 (-0.03, 0.60) | -0.06 (-0.38, 0.26) | -0.31 (-0.61, -0.02) * | -0.44 (-0.75, -0.14) * |
| **Transition** | -0.08 (-0.31, 0.14) | 0.27 (-0.25, 0.79) | 0.20 (-0.30, 0.69) | -0.07 (-0.58, 0.44) | -0.20 (-0.71, 0.30) | -0.53 (-1.03, -0.04) * |
| **Relaxed  border  restrictions** | 0.00 (-0.16, 0.16) | 0.59 (0.22, 0.97) * | 0.43 (0.07, 0.79) * | -0.09 (-0.46, 0.29) | -0.45 (-0.80, -0.10) * | -0.38 (-0.74, -0.02) * |

*All models are adjusted for covariates (age, sex, mental health status, employment status, and availability of COVID-19 vaccine).*

**P < 0.05*

**Table S3. Regression coefficients of the change in symptoms of depression (PHQ-2 score of 3 or more) from Circuit Breaker to each restriction period for each age group.**

|  | Coefficients (95% CI) | | | | | |
| --- | --- | --- | --- | --- | --- | --- |
|  | Change in Depression Symptoms (PHQ-2) for Ages: | | | | | |
|  | ***Full Population*** | **Ages 18 to 29** | **Ages 30 to 39** | **Ages 40 to 49** | **Ages 50 to 59** | **Ages Over 60** |
| **Circuit**  **Breaker** | *Reference* | *Reference* | *Reference* | *Reference* | *Reference* | *Reference* |
| **Phase**  **1 and 2** | 0.87 (0.79, 0.96) * | 0.94 (0.77, 1.14) | 0.90 (0.74, 1.10) | 0.86 (0.70, 1.06) | 0.75 (0.61, 0.93) * | 0.90 (0.69, 1.16) |
| **Phase 3** | 0.86 (0.77, 0.94) * | 1.02 (0.83, 1.25) | 1.00 (0.81, 1.22) | 0.84 (0.67, 1.04) | 0.67 (0.54, 0.84) * | 0.69 (0.53, 0.91) * |
| **Heightened**  **Alert** | 0.85 (0.77, 0.94) * | 1.01 (0.81, 1.26) | 0.89 (0.71, 1.10) | 0.86 (0.68, 1.08) | 0.78 (0.62, 0.99) * | 0.69 (0.52, 0.92) * |
| **Transition** | 0.80 (0.68, 0.96) * | 0.93 (0.66, 1.33) | 0.92 (0.65, 1.30) | 0.79 (0.54, 1.17) | 0.82 (0.55, 1.22) | 0.48 (0.28, 0.83) * |
| **Relaxed**  **Border**  **Restrictions** | 0.87 (0.77, 0.98) * | 1.13 (0.87, 1.45) | 1.05 (0.82, 1.34) | 0.86 (0.65, 1.13) | 0.64 (0.48, 0.85) * | 0.64 (0.45, 0.91) * |

*All models are adjusted for covariates (age, sex, mental health status, and employment status).*

**P < 0.05*

**Table S4. Regression coefficients of the change in symptoms of anxiety (GAD-2 score of 3 or more) from Circuit Breaker to each restriction period for each age group.**

|  | Coefficients (95% CI) | | | | | |
| --- | --- | --- | --- | --- | --- | --- |
|  | Change in Anxiety Symptoms (GAD-2) for Ages: | | | | | |
|  | ***Full Population*** | **Ages 18 to 29** | **Ages 30 to 39** | **Ages 40 to 49** | **Ages 50 to 59** | **Ages Over 60** |
| **Circuit**  **Breaker** | *Reference* | *Reference* | *Reference* | *Reference* | *Reference* | *Reference* |
| **Phase**  **1 and 2** | 0.98 (0.89, 1.08) | 1.18 (0.97, 1.44) | 1.23 (1.01, 1.50) * | 0.99 (0.80, 1.23) | 0.75 (0.61, 0.92) * | 0.71 (0.56, 0.92) * |
| **Phase 3** | 0.96 (0.87, 1.06) | 1.23 (1.00, 1.51) * | 1.31 (1.07, 1.62) * | 0.90 (0.72, 1.12) | 0.70 (0.56, 0.87) * | 0.65 (0.50, 0.85) * |
| **Heightened**  **Alert** | 0.97 (0.87, 1.08) | 1.25 (1.00, 1.56) * | 1.22 (0.98, 1.53) | 1.00 (0.79, 1.26) | 0.77 (0.61, 0.97) * | 0.62 (0.47, 0.81) * |
| **Transition** | 0.92 (0.78, 1.09) | 1.28 (0.90, 1.81) | 1.01 (0.71, 1.44) | 1.02 (0.70, 1.49) | 0.86 (0.58, 1.27) | 0.44 (0.26, 0.75) * |
| **Relaxed**  **Border**  **Restrictions** | 1.03 (0.91, 1.16) | 1.48 (1.15, 1.91) * | 1.29 (1.00, 1.66) | 1.03 (0.78, 1.36) | 0.79 (0.60, 1.03) | 0.60 (0.43, 0.83) * |

*All models are adjusted for covariates (age, sex, mental health status, and employment status).*

**P < 0.05*

**2. Results**

**2.1 Demographic Breakdowns**

Tables S6 and S7 describe the survey population breakdown for each of Singapore’s pandemic restriction periods. As there was a sharp increase in the number of participants preferring not to state whether they had been diagnosed with a mental health condition in later restriction periods compared to earlier ones (Table S6), the breakdowns of participants’ GAD-2 and PHQ-2 scores across pandemic waves were plotted (Figures S2 and S3 respectively). However, the proportion of participants with each GAD-2 and PHQ-2 score did not seem to significantly change in later waves compared to the initial ones.

**Table S6. Demographic breakdowns of survey population for each of the restriction periods during the study (N=34429).**

|  |  | **Population Subgroup Counts (and Percentages) by COVID Measure** | | | | | |
| --- | --- | --- | --- | --- | --- | --- | --- |
|  |  | **Circuit Breaker** | **Phase 1 and 2** | **Phase 3** | **Heightened**  **alert** | **Transition** | **Relaxed border restrictions** |
| Age group | 18–29 | 575 (20.29%) | 2560 (20.60%) | 1844 (20.40%) | 1255 (20.01%) | 191 (19.55%) | 574 (19.92%) |
|  | 30–39 | 617 (21.77%) | 2669 (21.48%) | 1956 (21.64%) | 1311 (20.91%) | 211 (21.60%) | 637 (22.11%) |
|  | 40–49 | 585 (20.64%) | 2573 (20.70%) | 1881 (20.81%) | 1215 (19.37%) | 197 (20.16%) | 555 (19.26%) |
|  | 50–59 | 594 (20.96%) | 2559 (20.59%) | 1840 (20.36%) | 1320 (21.05%) | 194 (19.86%) | 592 (20.55%) |
|  | 60+ | 463 (16.34%) | 2066 (16.63%) | 1518 (16.79%) | 1170 (18.66%) | 184 (18.83%) | 523 (18.15%) |
| Gender | Female | 1415 (49.93%) | 6244 (50.25%) | 4468 (49.43%) | 3130 (49.91%) | 484 (49.54%) | 1435 (49.81%) |
|  | Male | 1419 (50.07%) | 6183 (49.75%) | 4571 (50.57%) | 3141 (50.09%) | 493 (50.46%) | 1446 (50.19%) |
| Have mental health condition | No | 2722 (96.05%) | 11830 (95.20%) | 7299 (80.75%) | 4456 (71.06%) | 710 (72.67%) | 1979 (68.69%) |
|  | Prefer not  to say | 75 (2.65%) | 364 (2.93%) | 1579 (17.47%) | 1740 (27.75%) | 255 (26.10%) | 844 (29.30%) |
|  | Yes | 37 (1.31%) | 233 (1.87%) | 161 (1.78%) | 75 (1.20%) | 12 (1.23%) | 58 (2.01%) |
| Working status | Full time  employment | 1794 (63.30%) | 7882 (63.43%) | 5863 (64.86%) | 3995 (63.71%) | 619 (63.36%) | 1837 (63.76%) |
|  | Not working | 297 (10.48%) | 1372 (11.04%) | 974 (10.78%) | 731 (11.66%) | 125 (12.79%) | 366 (12.70%) |
|  | Other | 534 (18.84%) | 2428 (19.54%) | 1710 (18.92%) | 1206 (19.23%) | 175 (17.91%) | 519 (18.01%) |
|  | Unemployed | 209 (7.37%) | 745 (6.00%) | 492 (5.44%) | 339 (5.41%) | 58 (5.94%) | 159 (5.52%) |

**Table S7. Distribution of PHQ-4 Mean (SD) Scores by COVID restriction measure and demographic subgroups of survey population for each of Singapore’s restriction periods during the study (Mean=3.51, SD=3.27).**

|  |  | **Population Subgroup Mean PHQ-4 Scores (and SDs) by COVID Measure** | | | | | |
| --- | --- | --- | --- | --- | --- | --- | --- |
|  |  | **Circuit Breaker** | **Phase 1**  **and 2** | **Phase 3** | **Heightened**  **alert** | **Transition** | **Relaxed border restrictions** |
| Age group | 18–29 | 4.40 (3.02) | 4.65 (3.18) | 4.74 (3.21) | 4.78 (3.22) | 4.66 (3.02) | 5.01 (3.12) |
|  | 30–39 | 3.98 (3.10) | 4.06 (3.24) | 4.26 (3.24) | 4.20 (3.16) | 4.12 (3.13) | 4.36 (3.21) |
|  | 40–49 | 3.66 (3.38) | 3.51 (3.22) | 3.50 (3.20) | 3.57 (3.20) | 3.56 (3.19) | 3.58 (3.15) |
|  | 50–59 | 3.10 (3.30) | 2.83 (3.16) | 2.68 (3.07) | 2.88 (3.18) | 3.02 (3.37) | 2.81 (3.14) |
|  | 60+ | 2.46 (3.13) | 2.14 (2.98) | 1.93 (2.75) | 1.96 (2.88) | 1.85 (2.60) | 2.03 (2.85) |
| Gender | Female | 3.65 (3.21) | 3.51 (3.22) | 3.52 (3.23) | 3.44 (3.18) | 3.39 (3.13) | 3.62 (3.22) |
|  | Male | 3.48 (3.30) | 3.49 (3.33) | 3.46 (3.30) | 3.55 (3.39) | 3.55 (3.30) | 3.58 (3.33) |
| Have mental health condition | No | 3.53 (3.24) | 3.43 (3.24) | 3.34 (3.21) | 3.23 (3.23) | 3.22 (3.15) | 3.34 (3.25) |
|  | Prefer not  to say | 3.79 (3.08) | 3.80 (3.30) | 3.89 (3.31) | 4.10 (3.28) | 3.97 (3.17) | 4.12 (3.24) |
|  | Yes | 5.73 (3.96) | 6.49 (3.61) | 6.10 (3.88) | 5.52 (3.92) | 7.25 (4.61) | 4.90 (3.38) |
| Working status | Full time  employment | 3.61 (3.23) | 3.56 (3.19) | 3.57 (3.19) | 3.63 (3.22) | 3.57 (3.21) | 3.81 (3.23) |
|  | Not working | 2.45 (3.00) | 2.29 (3.02) | 2.06 (2.88) | 2.06 (2.92) | 2.66 (3.19) | 2.05 (2.94) |
|  | Other | 3.65 (3.23) | 3.63 (3.30) | 3.71 (3.31) | 3.61 (3.37) | 3.33 (3.03) | 3.79 (3.31) |
|  | Unemployed | 4.58 (3.45) | 4.64 (3.88) | 4.59 (3.79) | 4.62 (3.59) | 4.53 (3.54) | 4.08 (3.40) |

**Figure S2. Breakdown of participants’ GAD-2 scores across Singapore’s restriction periods.**

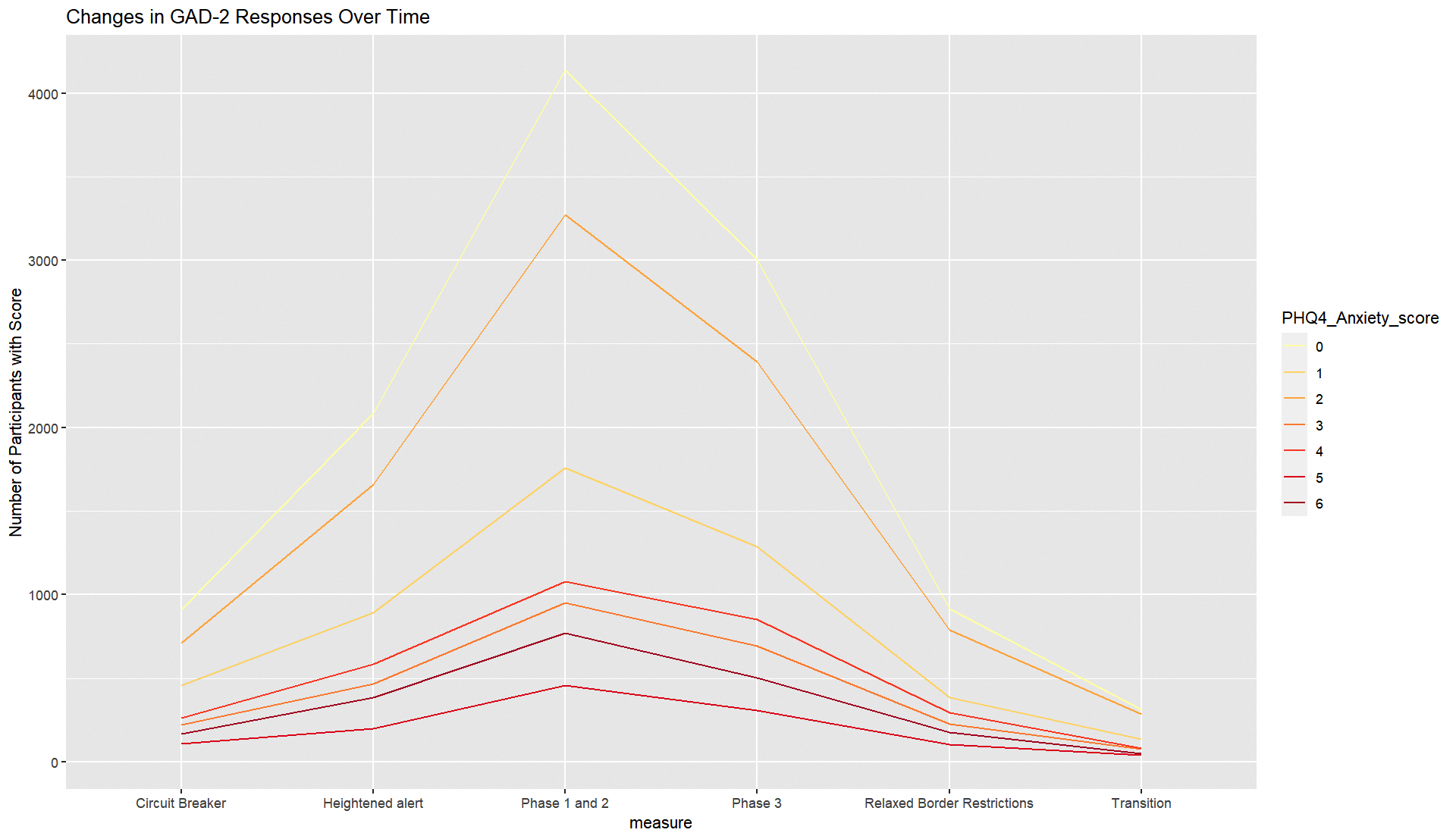

**Figure S3. Breakdown of participants’ PHQ-2 scores across Singapore’s restriction periods**

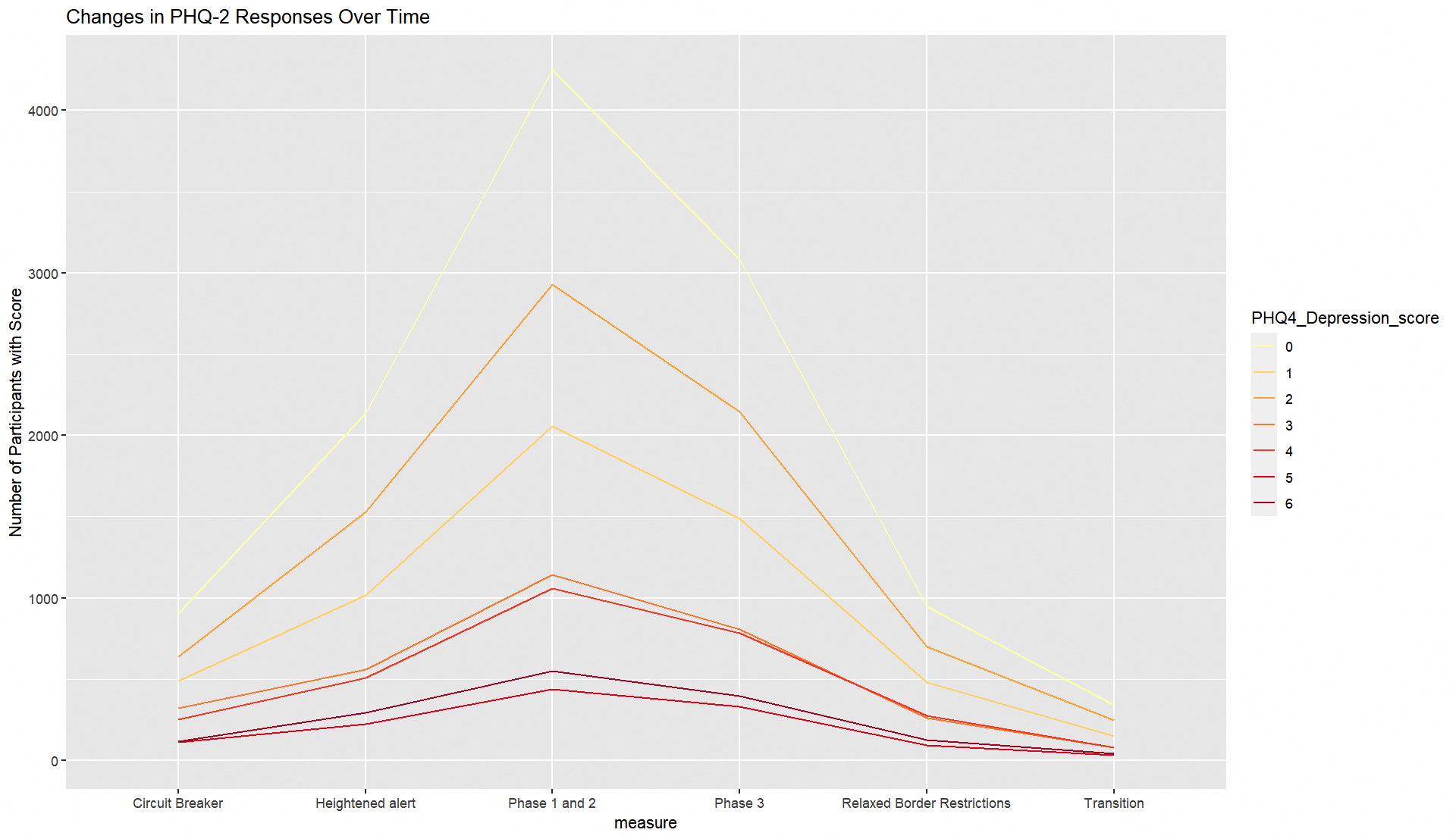

**2.2. Additional Regressions**

**Table S8. Multiple linear regression of the change in psychological distress (total PHQ-4 score) from Circuit Breaker to each restriction period for different demographic groups.**

|  | Coefficients (95% CI) | | | | |
| --- | --- | --- | --- | --- | --- |
|  | Change in Anxiety and Depression symptoms (PHQ-4) from Circuit Breaker to: | | | | |
|  | **Phase 1 and 2** | **Phase 3** | **Heightened alert** | **Transition** | **Relaxed Border**  **Restrictions** |
| *Gender* |  |  |  |  |  |
| Females | -0.14 (-0.32, 0.03) | -0.14 (-0.34, 0.06) | -0.21 (-0.40, -0.02) * | -0.27 (-0.58, 0.04) | -0.08 (-0.31, 0.14) |
| Males | 0.02 (-0.16, 0.21) | 0.01 (-0.20, 0.22) | 0.13 (-0.07, 0.33) | 0.11 (-0.22, 0.44) | 0.09 (-0.15, 0.33) |
| *Employment status* | | | | | |
| Employed full time | -0.07 (-0.23, 0.08) | -0.05 (-0.23, 0.13) | 0.01 (-0.17, 0.18) | -0.08 (-0.36, 0.20) | 0.11 (-0.09, 0.31) |
| Not working | -0.15 (-0.50, 0.20) | -0.25 (-0.65, 0.16) | -0.40 (-0.79, -0.02) * | 0.19 (-0.41, 0.80) | -0.38 (-0.82, 0.06) |
| Unemployed | -0.00 (-0.56, 0.56) | 0.20 (-0.46, 0.87) | 0.04 (-0.60, 0.68) | -0.03 (-1.09, 1.03) | -0.47 (-1.23, 0.28) |
| Other | -0.00 (-0.29, 0.29) | -0.07 (-0.40, 0.26) | -0.04 (-0.36, 0.28) | -0.35 (-0.88, 0.19) | 0.04 (-0.34, 0.42) |
| *Mental health condition* | | | | | |
| Have mental health condition | 0.38 (-0.86, 1.61) | 0.25 (-1.14, 1.64) | -0.36 (-1.76, 1.05) | 2.08 (-0.27, 4.43) | -1.04 (-2.51, 0.43) |
| No mental health condition | -0.07 (-0.20, 0.06) | -0.04 (-0.20, 0.11) | -0.07 (-0.22, 0.08) | -0.07 (-0.32, 0.18) | 0.01 (-0.17, 0.19) |
| Prefer not to disclose mental health condition | -0.30 (-1.08, 0.49) | -0.26 (-1.00, 0.49) | -0.07 (-0.81, 0.66) | -0.30 (-1.12, 0.51) | -0.05 (-0.80, 0.70) |

*All models are adjusted for covariates (age, sex, mental health status, employment status, and availability of COVID-19 vaccine).*

**P < 0.05*

**Table S9. Univariate and multiple linear regression of the change in psychological distress (total PHQ-4 score) from Circuit Breaker to each restriction period for the full population.**

|  | **Coefficients (95% CI)** | |
| --- | --- | --- |
|  | **Change in psychological distress (PHQ-4) for whole population using:** | |
|  | **Univariate regression** | **Multivariate regression** |
| **Circuit Breaker** | *Reference* | *Reference* |
| **Phase 1 and 2** | -0.06 (-0.19, 0.08) | -0.06 (-0.19, 0.06) |
| **Phase 3** | -0.09 (-0.22, 0.05) | -0.06 (-0.21, 0.08) |
| **Heightened alert** | -0.05 (-0.19, 0.10) | -0.05 (-0.19, 0.09) |
| **Transition** | -0.06 (-0.30, 0.18) | -0.08 (-0.31, 0.14) |
| **Relaxed Border Restrictions** | 0.04 (-0.13, 0.21) | 0.00 (-0.16, 0.16) |

*All models are adjusted for covariates (age, sex, mental health status, employment status, and availability of COVID-19 vaccine).*

**P < 0.05*

**2.2. Baseline Comparisons**

**Table S10. Demographic breakdown of survey population for Singapore’s Circuit Breaker**

|  | **Sample characteristics** | **PHQ4 score** |
| --- | --- | --- |
|  | **Count (%)** | **M (SD)** |
| *Full population* | *2834 (100.00%)* | *3.57 (3.25)* |
| 18–29 | 575 (20.29%) | 4.40 (3.02) |
| 30–39 | 617 (21.77%) | 3.98 (3.10) |
| 40–49 | 585 (20.64%) | 3.66 (3.38) |
| 50–59 | 594 (20.96%) | 3.10 (3.30) |
| 60+ | 463 (16.34%) | 2.46 (3.13) |

**Table S11. Multiple linear regression of the difference in psychological distress between 40 to 49 year-olds (reference group) and every other age group during Circuit Breaker.**

|  | **Coefficient (95% CI)** |
| --- | --- |
| **Age group** | **Difference in PHQ4 score compared to 40- to 49-year-olds** |
| 40–49 | Reference |
| 18–29 | 0.74 (0.36, 1.11) |
| 30–39 | 0.30 (-0.06, 0.66) |
| 50–59 | -0.53 (-0.89, -0.16) |
| 60+ | -1.02 (-1.43, -0.61) |

*Model is adjusted for covariates (sex, mental health status, employment status).*

**P < 0.05*
